# Clinical Diversity and Outcomes of Progressive Familial Intrahepatic Cholestasis Diagnosed by Whole Genome Sequencing in Pakistani Children

**DOI:** 10.1101/2024.02.26.24303272

**Authors:** Huma Arshad Cheema, Aliaksandr Skrahin, Anjum Saeed, Zafar Fayyaz, Muhammad Arshad Alvi, Muhammad Nadeem Anjum, Nadia Waheed, Khalil Ur Rehman, Ahmad Malik, Volha Skrahina, Arndt Rolfs

## Abstract

Progressive familial intrahepatic cholestasis (PFIC) is a rare group of genetic disorders that typically present in infants and children, often progressing to end-stage liver disease. Using whole genome sequencing (WGS) for diagnosis, we aimed to assess phenotypic features and outcomes, in Pakistani children with different types of PFIC. The study included 116 pediatric participants with five PFIC types: PFIC1, *ATP8B1* gene (n=19); PFIC2, *ABCB11* (n=28); PFIC3, *ABCB4* (n=52); PFIC4 *TJP2* (n=15); and PFIC5 *NR1H4* (n=2). Seventy unique variants were identified across the five genes. Age at genetic diagnosis was higher in PFIC3 patients. Clinical and laboratory findings showed significant overlap among all PFIC types. PFIC3 had a less aggressive course and better survival outcomes compared to PFIC1, PFIC2, and PFIC4. The cumulative survival rate was significantly higher at 89% (95% CI 43-98%) for patients who underwent liver transplantation, compared to 9% (95% CI 1-29%) for those who did not (p=0.016). The study provides the first comprehensive analysis of PFIC in Pakistani children, highlighting significant clinical overlap and the critical need for early genetic diagnosis using WGS. The findings underscore the importance of personalized treatment approaches, including early consideration for liver transplantation, to improve patient outcomes.

## 1. Introduction

Genetic factors are implicated in a significant proportion of pediatric cholestatic liver diseases, with estimates suggesting that at least 45% have a genetic basis [1]. Progressive familial intrahepatic cholestasis (PFIC) is a rare group of autosomal recessive inherited disorders characterized by intrahepatic cholestasis, presenting with jaundice, pruritus, and failure to thrive in infants and children, typically progressing to end-stage liver disease [1–3]. In the Online Mendelian Inheritance in Man (OMIM) database (https://www.omim.org/), bi-allelic (homozygous or compound heterozygous) variants in 12 genes have been associated with PFIC, including *ATP8B1, ABCB11, ABCB4, TJP2, NR1H4, SLC51A, USP53, KIF12, ZFYVE19, MYO5B, SEMA7A*, and *VPS33B* [4]. Worldwide, PFIC has an estimated incidence of 1 per 50,000-100,000 live births [5]; however, the exact incidence in Pakistan is unknown.

Clinical presentations and laboratory findings in PFIC patients often overlap between various PFIC types. While it is possible to suspect and differentiate between different PFIC types based on clinical presentation and basic diagnostic tests, confirmatory diagnosis usually relies on genetic testing. Whole genome sequencing (WGS) in routine diagnostics for patients with cholestatic liver disease has been implemented in Pakistani hospitals. This study aims to assess the clinical and laboratory diversity, as well as the overlap in clinical and laboratory diagnostic findings, among various types of PFIC in a large cohort of Pakistani children at the time of their genetic confirmation. The study also evaluates the clinical outcomes of the PFIC patients.

## 2. Results

### 2.1. Participants

Of all children with clinical cholestasis tested with WGS in Pakistani hospitals during the period from January 2019 to January 2023, 116 patients with genetically confirmed PFIC were included in our study. There were patients with five different types of PFIC: PFIC1 (*ATP8B1*) - 19 (16.4%), PFIC2 (*ABCB11*) - 28 (24.1%), PFIC3 (*ABCB4*) - 52 (44.8%), PFIC4 (*TJP2*) - 15 (12.9%), PFIC5 (*NR1H4*) - 2 (1.7%).

### 2.2. PFIC variants

Seventy unique PFIC variants were identified in the study (Table S1). Some of these variants were found in multiple patients, with the most common variant occurring in up to 10 patients. In each PFIC type, cases were marked by homozygous variants in their respective genes (*ATP8B1, ABCB11, ABCB4, TJP2*, and *NR1H4*), all classified as either pathogenic or likely pathogenic (P/LP). One rare occurrence of PFIC2 involved a compound heterozygous mutation in the *ABCB11* gene.

Additionally, a singular case of PFIC3, associated with the *ABCB4* gene, exhibited a variant of uncertain significance (VUS). In terms of impact on the protein, our study detected the following types of variants: frameshift - 15 variants in 25 patients; nonsense - 15 variants in 30 patients; stop gain - 2 variants in 3 patients; splice - 4 variants in 5 patients; start loss - 1 variant in 1 patient; missense - 30 variants in 50 patients; and substitution - 2 variants in 2 patients. In 64 patients, 37 unique variants affecting protein function were classified as truncated.

### 2.3. Clinical and laboratory findings

Detailed clinical and laboratory findings pertaining to these cases are outlined in Table 1, Table 2 and Table 3. Only two male participants diagnosed with PFIC5 were included in the study. Given the small number of PFIC5 patients, their comparisons with other groups were omitted.

**Table 1.**
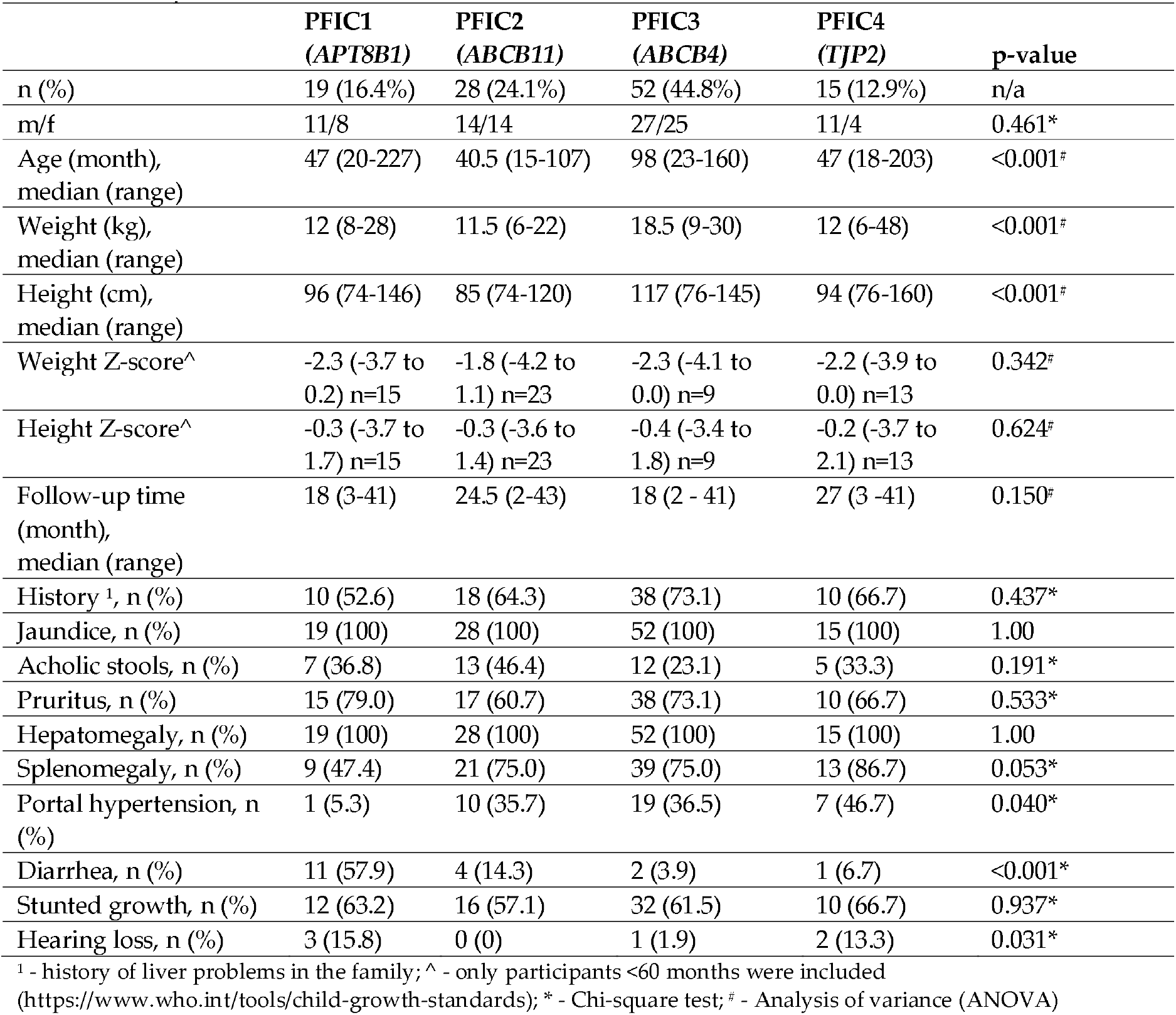
PFIC patients’ clinical data.

|  | PFIC1<br>(*APT8B1*) | PFIC2<br>(*ABCB11*) | PFIC3<br>(*ABCB4*) | PFIC4<br>(*TJP2*) | p-value |
| --- | --- | --- | --- | --- | --- |
| n (%) | 19 (16.4%) | 28 (24.1%) | 52 (44.8%) | 15 (12.9%) | n/a |
| m/f | 11/8 | 14/14 | 27/25 | 11/4 | 0.461* |
| Age (month),<br>median (range) | 47 (20-227) | 40.5 (15-107) | 98 (23-160) | 47 (18-203) | <0.001<sup>‡</sup> |
| Weight (kg),<br>median (range) | 12 (8-28) | 11.5 (6-22) | 18.5 (9-30) | 12 (6-48) | <0.001<sup>‡</sup> |
| Height (cm),<br>median (range) | 96 (74-146) | 85 (74-120) | 117 (76-145) | 94 (76-160) | <0.001<sup>‡</sup> |
| Weight Z-score<sup>^</sup> | -2.3 (-3.7 to<br>0.2) n=15 | -1.8 (-4.2 to<br>1.1) n=23 | -2.3 (-4.1 to<br>0.0) n=9 | -2.2 (-3.9 to<br>0.0) n=13 | 0.342<sup>‡</sup> |
| Height Z-score<sup>^</sup> | -0.3 (-3.7 to<br>1.7) n=15 | -0.3 (-3.6 to<br>1.4) n=23 | -0.4 (-3.4 to<br>1.8) n=9 | -0.2 (-3.7 to<br>2.1) n=13 | 0.624<sup>‡</sup> |
| Follow-up time<br>(month),<br>median (range) | 18 (3-41) | 24.5 (2-43) | 18 (2 - 41) | 27 (3 -41) | 0.150<sup>‡</sup> |
| History <sup>1</sup>, n (%) | 10 (52.6) | 18 (64.3) | 38 (73.1) | 10 (66.7) | 0.437* |
| Jaundice, n (%) | 19 (100) | 28 (100) | 52 (100) | 15 (100) | 1.00 |
| Acholic stools, n (%) | 7 (36.8) | 13 (46.4) | 12 (23.1) | 5 (33.3) | 0.191* |
| Pruritus, n (%) | 15 (79.0) | 17 (60.7) | 38 (73.1) | 10 (66.7) | 0.533* |
| Hepatomegaly, n (%) | 19 (100) | 28 (100) | 52 (100) | 15 (100) | 1.00 |
| Splenomegaly, n (%) | 9 (47.4) | 21 (75.0) | 39 (75.0) | 13 (86.7) | 0.053* |
| Portal hypertension, n<br>(%) | 1 (5.3) | 10 (35.7) | 19 (36.5) | 7 (46.7) | 0.040* |
| Diarrhea, n (%) | 11 (57.9) | 4 (14.3) | 2 (3.9) | 1 (6.7) | <0.001* |
| Stunted growth, n (%) | 12 (63.2) | 16 (57.1) | 32 (61.5) | 10 (66.7) | 0.937* |
| Hearing loss, n (%) | 3 (15.8) | 0 (0) | 1 (1.9) | 2 (13.3) | 0.031* |

<sup>1</sup> - history of liver problems in the family; <sup>^</sup> - only participants <60 months were included
(https://www.who.int/tools/child-growth-standards); \* - Chi-square test; <sup>‡</sup> - Analysis of variance (ANOVA)

**Table 2.** PFIC patients’ laboratory findings: number of patients with laboratory values higher than the upper limit of normal (ULN) or lower than the lower limit of normal (LLN).

| Test | PFIC1 n=19 | PFIC2 n=28 | PFIC3 n=52 | PFIC4 n=15 | p-value* |
| --- | --- | --- | --- | --- | --- |
| Bil. tot. > 1.0 mg/dL | 19 (100%) | 28 (100%) | 51 (98.1%) | 15 (100%) | 1.00 |
| Bil. con. > 0.3 mg/dL | 19 (100%) | 28 (100%) | 52 (100%) | 15 (100%) | 1.00 |
| GGT > 55 IU | 13 (68.4%) | 12 (42.9%) | 50 (96.2%) | 6 (40%) | <0.001 |
| ALP > 350 IU | 2 (10.5%) | 5 (17.9%) | 22 (42.3%) | 6 (40%) | 0.001 |
| ALT > 40 IU | 18 (94.7%) | 27 (96.4%) | 52 (100%) | 15 (100%) | 0.378 |
| AST > 40 IU | 19 (100%) | 27 (96.4%) | 51 (98.1%) | 15 (100%) | 0.765 |
| aPTT(R) > 1.2 | 19 (100%) | 28 (100%) | 49 (94.2%) | 15 (100%) | 0.565 |
| INR > 1.2 | 18 (94.7%) | 24 (85.7%) | 44 (84.6%) | 13 (86.7%) | 0.731 |
| PLT < 150*10<sup>9</sup> /L | 1 (5.3%) | 2 (7.1%) | 9 (17.3%) | 3 (20.0%) | 0.345 |
| Hg < 110 g/L | 16 (84.2%) | 25 (89.3%) | 44 (84.6%) | 13 (86.7%) | 0.943 |

Bil tot - bilirubin total; Bil. con. - bilirubin conjugated; GGT - Gamma-glutamyl transferase; ALP - alkaline phosphatase; AST - aspartate aminotransferase; ALT - alanine aminotransferase; aPTT(R) - activated partial thromboplastin time (ratio); INR - international normalized ratio; PLT - blood platelet count; Hb - hemoglobin; AFT - alpha-fetoprotein; n/a - not available. \* - Chi-square test

**Table 3.**
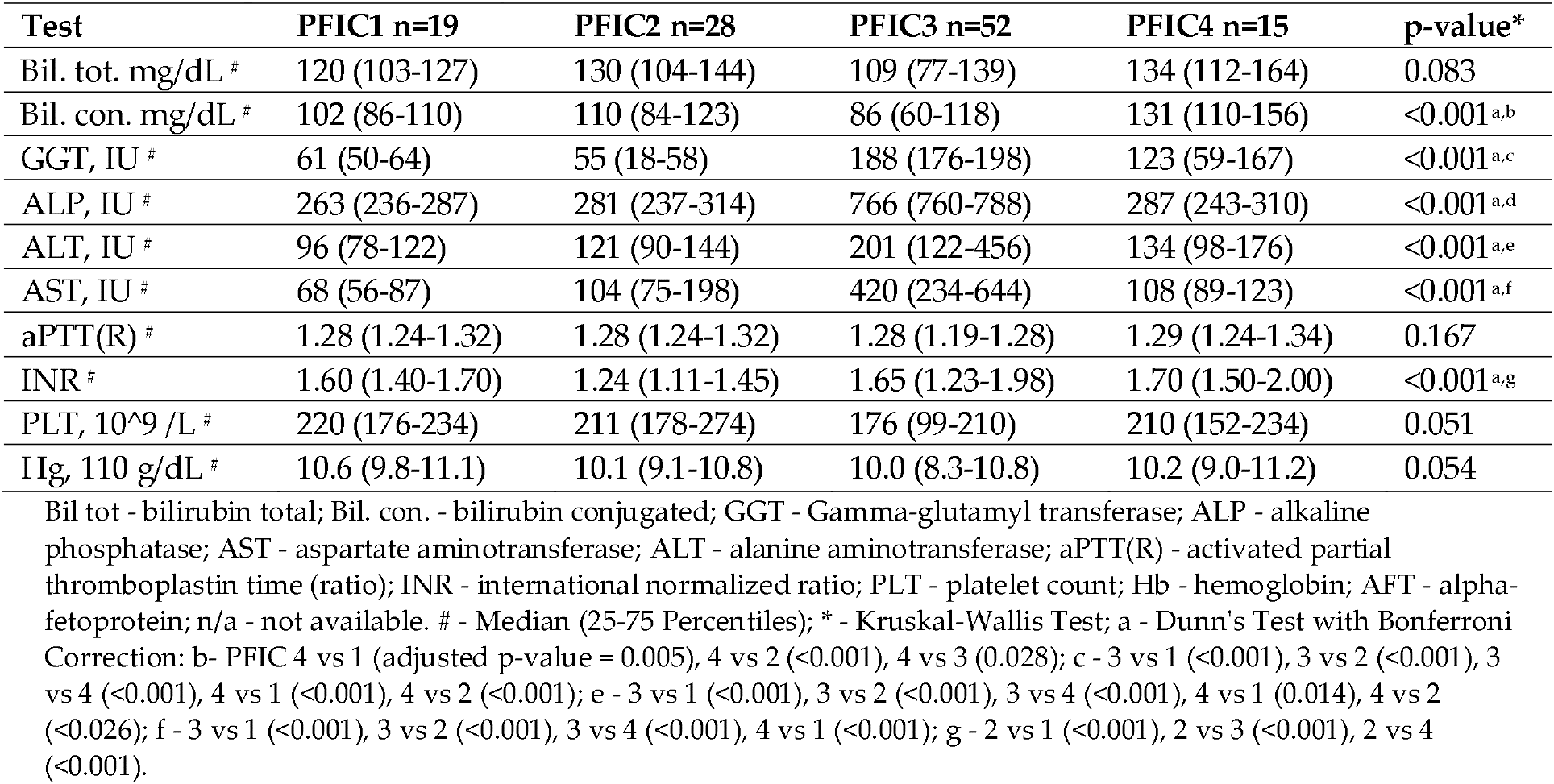
PFIC patients’ laboratory values.

| Test | PFIC1 n=19 | PFIC2 n=28 | PFIC3 n=52 | PFIC4 n=15 | p-value* |
| --- | --- | --- | --- | --- | --- |
| Bil. tot. mg/dL # | 120 (103-127) | 130 (104-144) | 109 (77-139) | 134 (112-164) | 0.083 |
| Bil. con. mg/dL # | 102 (86-110) | 110 (84-123) | 86 (60-118) | 131 (110-156) | <0.001<sup>a,b</sup> |
| GGT, IU # | 61 (50-64) | 55 (18-58) | 188 (176-198) | 123 (59-167) | <0.001<sup>a,c</sup> |
| ALP, IU # | 263 (236-287) | 281 (237-314) | 766 (760-788) | 287 (243-310) | <0.001<sup>a,d</sup> |
| ALT, IU # | 96 (78-122) | 121 (90-144) | 201 (122-456) | 134 (98-176) | <0.001<sup>a,e</sup> |
| AST, IU # | 68 (56-87) | 104 (75-198) | 420 (234-644) | 108 (89-123) | <0.001<sup>a,f</sup> |
| aPTT(R) # | 1.28 (1.24-1.32) | 1.28 (1.24-1.32) | 1.28 (1.19-1.28) | 1.29 (1.24-1.34) | 0.167 |
| INR # | 1.60 (1.40-1.70) | 1.24 (1.11-1.45) | 1.65 (1.23-1.98) | 1.70 (1.50-2.00) | <0.001<sup>a,g</sup> |
| PLT, 10<sup>9</sup> /L # | 220 (176-234) | 211 (178-274) | 176 (99-210) | 210 (152-234) | 0.051 |
| Hg, 110 g/dL # | 10.6 (9.8-11.1) | 10.1 (9.1-10.8) | 10.0 (8.3-10.8) | 10.2 (9.0-11.2) | 0.054 |

Bil tot - bilirubin total; Bil. con. - bilirubin conjugated; GGT - Gamma-glutamyl transferase; ALP - alkaline phosphatase; AST - aspartate aminotransferase; ALT - alanine aminotransferase; aPTT(R) - activated partial thromboplastin time (ratio); INR - international normalized ratio; PLT - platelet count; Hb - hemoglobin; AFT - alpha-fetoprotein; n/a - not available. # - Median (25-75 Percentiles); \* - Kruskal-Wallis Test; a - Dunn's Test with Bonferroni Correction: b- PFIC 4 vs 1 (adjusted p-value = 0.005), 4 vs 2 (<0.001), 4 vs 3 (0.028); c - 3 vs 1 (<0.001), 3 vs 2 (<0.001), 3 vs 4 (<0.001), 4 vs 1 (<0.001), 4 vs 2 (<0.001); e - 3 vs 1 (<0.001), 3 vs 2 (<0.001), 3 vs 4 (<0.001), 4 vs 1 (0.014), 4 vs 2 (<0.026); f - 3 vs 1 (<0.001), 3 vs 2 (<0.001), 3 vs 4 (<0.001), 4 vs 1 (<0.001); g - 2 vs 1 (<0.001), 2 vs 3 (<0.001), 2 vs 4 (<0.001).

When comparing PFIC1, 2, 3, and 4 groups, at the time of the genetic diagnosis, both genders were equally represented across all groups (Table 1). Patients diagnosed with PFIC3 were older than those in other groups (p-value <0.001). As a result, PFIC3 patients had significantly higher weight and height measurements compared to other groups (p-value <0.001). However, when assessing growth relative to age and gender norms using height and weight z-scores [6], there were no significant differences between the groups. The follow-up period ranged from 2 to 43 months, with clinical manifestations varying from mild/moderate to end-stage liver disease. A significant proportion (52.6-73.1%) of patients in each group reported a family history of liver issues. Common clinical findings included hepatomegaly and jaundice (incl. scleral icterus), with increased direct and total bilirubin levels observed in all participants. Interestingly (Table 2), the most pronounced increase in conjugated bilirubin was observed in patients with PFIC4 (Table 3). Acholic stool was more common in PFIC2 patients (46.4%) and less so in PFIC3 (23.1%), though not statistically significant. Pruritus occurred less frequently in PFIC2 (60.7%) compared to other groups (66.7%-79.0%), also without statistical significance. The diagnosis of pruritus in preverbal children was based on clinical judgment, taking into account several key factors: observable physical signs such as scratches, lichenification, and secondary skin infections; behavioral indicators including excessive rubbing, agitation or irritability, and disturbed sleep patterns; and the child’s response to antipruritic treatment. Splenomegaly was less common in PFIC1 patients (47.4%) compared to others (75.0-86.7%), though not statistically significant. Portal hypertension syndrome, marked by symptoms like esophageal varices, ascites, caput medusae, encephalopathy, and leg swelling, was significantly less common in PFIC1 patients (5.3%) compared to other groups (35.7-46.7%), with a p-value of 0.040. The growth of the participants was evaluated using the height-for-age and weight-to-age metric – Z-scores. Stunting was identified when the Z-score value fell below two standard deviations (SD) from the median of the World Health Organization (WHO) Child Growth Standards (https://www.who.int/tools/child-growth-standards) [6]. The stunted growth was observed in more than half of the participants in all groups (57.1-66.7%). Early diagnosis of hearing loss is essential, as it significantly influences language development, social skills, and overall learning outcomes. In our study, the otoacoustic emissions (OAE) test [7] was employed to identify hearing loss, with six patients (PFIC1 -3; PFIC3 – 1, and PFIC4 – 2) subsequently diagnosed with the condition. The PFIC1 and PFIC4 patients were more prone (p-value=0.031) to develop hearing loss. Almost all patients (84.6-100%) exhibited abnormal blood coagulation tests (activated Partial Thromboplastin Time / Ratio [aPTT/R]>1.2 and/or International Normalized Ratio [INR]>1.2) (Table 2), the INR was least elevated in PFIC2 patients (Table 3). Diarrhea was reported in the majority (59.4%) of PFIC1 patients (p-value<0.001), with lower frequencies in other groups (3.9-14.3%).

Gamma-glutamyl transferase (GGT) levels were elevated in one-third of PFIC1 (68.4%), about half of PFIC2 (42.9%) and PFIC4 (40.0%) patients, and in almost all (96.2%) of PFIC3 patients (p-value<0.001) (Table 2). The increase in GGT was most pronounced in patients with PFIC3 (p-value < 0.001), where levels were more than three times the upper limit of normal (3ULN); for patients with PFIC4, GGT levels were higher than 2ULN. In contrast, the increase in GGT levels for patients with PFIC1 and PFIC2 did not exceed 2ULN (Table 3). Almost all patients in each group showed elevated aspartate aminotransferase (AST) and alanine aminotransferase (ALT) levels (Table 2) with the most pronounced increase in PFIC3 patients (Table 3). The elevation (Table 3) and prevalence (Table 2) of alkaline phosphatase (ALP) were also highest in PFIC3 patients. Alpha-fetoprotein (AFP) levels, were tested in 7 (2 - PFIC2, 3 - PFIC3, and 1 - PFIC4) patients during the period of treatment and follow-up. Moderate elevations (<400 mcg/L) were revealed in 2/2 PFIC2, 2/3 in PFIC3 and 1/1 PFIC4 patients. Only a small proportion of patients (5.3-20.0%) exhibited low platelet counts, with no differences between the groups. Anemia was typical for the vast majority (84-89%) of patients in all groups. Since the study participants underwent WGS, other common genetic causes of anemia such as sideroblastic anemia, Fanconi anemia, and sickle cell disease were effectively ruled out.

The two male participants, diagnosed with PFIC5 within the age range of 0-5 years (not included in the Tables), consistently exhibited jaundice, scleral icterus, hepatomegaly, splenomegaly, and stunted growth at the time of genetic diagnosis. However, neither showed signs of acholic stools or diarrhea. One patient displayed symptoms of pruritus. They demonstrated elevated levels of both total and conjugated bilirubin. Their GGT and ALP levels remained within normal ranges. However, there were elevations in ALT and AST, 121 IU, 123 IU, and 86I U, 87 IU, respectively. Blood coagulation was also affected: aPTT/R - 1.32 and 1.34, and INR - 1.54 and 1.62, accordingly. Both patients maintained normal hemoglobin levels and platelet counts. Their follow-up periods were 18 and 26 months, respectively. Both participants died.

### 2.4. Management and outcomes

In our cohort, the treatment strategy for the patients primarily included oral administration of ursodeoxycholic acid (UDCA) and fat-soluble vitamins (A, D, E, K), supplemented by nutritional interventions. For symptomatic relief of pruritus, agents such as cholestyramine and rifampicin were prescribed. Surgical interventions were also part of the treatment regimen, with nine patients undergoing Biliary Diversion (BD) surgery and ten patients receiving Liver Transplant (LT). Recognizing the hereditary nature of PFICs, and the frequent delays in generic diagnosis and treatment initiation, we focused on assessing cumulative survival rather than post-diagnostic (on-treatment) survival. Over 20 years (240 months), the cumulative survival rate for all PFIC patients was 20% (95% CI 5-41%) (Figure 1A). Of the 10 PFIC patients who underwent LT, only 1 patient died, in contrast to 44 of the 106 patients (42%) who were treated with standard methods. Despite LT being performed in cases with severe disease and/or at an advanced stage, the cumulative survival rate of patients with LT was significantly higher at 89% (95% CI 43-98%), compared to 9% (95% CI 1-29%) for those not undergoing LT, p-value = 0.016 (Figure 1B). Among the four PFIC types, PFIC3 exhibited a less aggressive course, with a better survival outcome compared to PFIC1, PFIC2, and PFIC4, p-value = 0.023 (Figure 1C). Cumulative survival of patients with non-truncated genetic variants was higher, 32% (95% CI 8-60%) (not significantly), than in patients with truncated variants, 14% (95% CI 1-41%) (Figure 1D).

**Figure 1.**
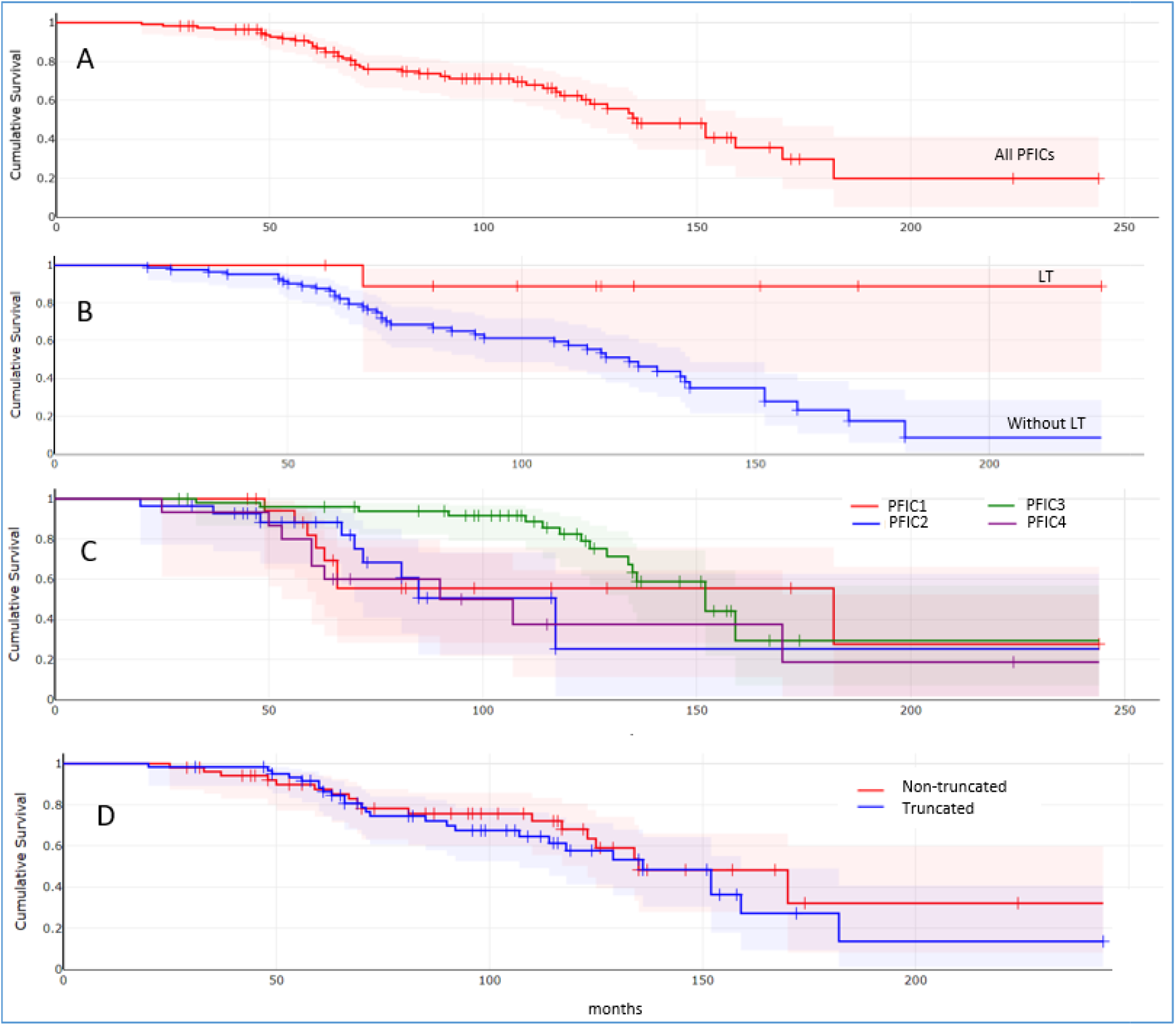
PFIC patients’ cumulative survival (95% Confidence Interval [CI]). **A:** all PFIC patients’ cumulative survival - 20% (95% CI 5-41%); B: cumulative survival with liver transplant (LT) 89% - (95% CI 43-98%) vs without liver transplant - 9% (95% CI 1-29%), p-value = 0.016; C: cumulative survival of different PFICs: PFIC1 - 27% (95% CI 2-66%), PFIC2 - 25% (95% CI 2-63%); PFIC3 - 29% (95% CI 7-57%); PFIC4 - 19% (95% CI 1-52%). PFIC3 showed better survival outcome, p-value = 0.023; D: cumulative survival of patients with truncated variants - 14% (95% CI 1-41%) vs non-truncated variants - 32% (95% CI 8-60%), p-value=0.547.

Specifically, for PFIC1 (ATP8B1), alongside medical treatments, 3 of 19 patients underwent BD, and 5 received LT. The 12-year survival rate was 27% (95% CI 2-66%); without LT, this rate decreased to 18% (95% CI 1-53%), p-value = 0.194. In the PFIC2 (*ABCB11*) group, five underwent BD and one received LT. The 20-year survival was 25% (95% CI 2-63%) (Figure 1C). Notably, none of the six patients who underwent BD or LT died, in contrast to 9 of 22 patients (41%) on standard medical treatment. For PFIC3 (*ABCB4*), three patients received LT. None of the PFIC3 LT patients died, while 16 (31%) of the other PFIC3 patients did. In the PFIC4 (*TJP2*) patients, the cumulative survival rate was 19% (95% CI 1-52%) (Figure 1C). One patient who underwent LT is currently alive. In contrast, 9 out of the remaining 14 patients (60%), have died. Both patients with PFIC5 (*NR1H4*) experienced severe progressive disease and passed away having not undergone any surgical interventions.

## 3. Discussion

In our study, we aimed to determine the distribution, disease characteristics, and clinical outcomes of patients with PFICs in Pakistan. In our cohort, PFIC3 emerges as the predominant type, representing nearly half of the cases (44.8%). This is followed by PFIC2, PFIC1, PFIC4, and PFIC5, which comprise 24.1%, 16.4%, 12.9%, and 1.7% of the cases, respectively. Earlier studies included diverse patient populations from France [8], Germany [9], India [10], the United States (US), without specifying the ethnicity [11], and the United States, encompassing Caucasian, African-American, Japanese, and Korean origins [12]. In these studies, PFIC1 and PFIC2 were the most frequent types, followed by PFIC3; PFIC4 and PFIC5 were much rarer. However, PFIC3 was the most common type (59.5%) in the study from Saudi Arabia followed by PFIC2 (34.2%), PFIC1 (5.1%), and PFIC4 (1.3%) [5].

Clinical and laboratory manifestations, as well as the disease course of various PFIC types, have been extensively documented in a multitude of studies, ranging from individual case reports to comprehensive reviews collating data from numerous publications on PFICs. The clinical presentations and laboratory findings in our study largely align with those reported previously. However, we have identified some particular features that distinguish our findings.

Generally, PFIC1 and PFIC2 typically manifest within the first year of life, whereas PFIC3 presents a broader age range from infancy to early adulthood; PFIC4 and PFIC5 usually exhibit symptoms in early childhood, although the exact onset can vary due to the rarity and variability of these types [2,5,13,14]. In our study, at the time of genetic diagnosis, PFIC3 patients were the oldest (median – 98 months) compared to other PFIC types diagnosed within 40-48 months of age (p-value < 0.001). This finding aligns with previous studies. However, our results suggest that PFIC genetic diagnosis in Pakistan may be delayed by several months, highlighting the need for increased awareness and timely diagnostic protocols to improve early detection and management.

In our study, the majority of patients in all PFIC groups reported a family history of liver diseases. Almost all PFIC variants, with only one exception, were homozygous rather than compound heterozygous. This suggests a highly consanguineous population within our cohort. Generally, consanguineous marriages in Pakistan are reported to range from 46–98%, depending on the region, which correlates with high rates of genetically inherited diseases and infant mortality [15].

Jaundice, due to elevated conjugated bilirubin, is present in all PFIC patients. Conjugated bilirubin was most elevated in PFIC4 and less elevated in PFIC3 with a significant difference. This disparity is likely due to the underlying genetic mutations and their impact on bile secretion pathways. PFIC4, caused by mutations in the TJP2 gene, results in severe disruption of bile canalicular membrane integrity, leading to higher conjugated bilirubin levels [16–18]. In contrast, PFIC3, associated with mutations in the ABCB4 gene, affects phospholipid transport, resulting in relatively less conjugated bilirubin elevation [16,19,20].

Acholic stool is a common cholestatic symptom. The proportion of patients with PFICs affected and the severity of this symptom can vary depending on bile secretion and flow [2,21]. In our study, acholic stool was observed in 23.1% to 46.4% of patients across different PFIC types, without a statistically significant difference between the groups.

Pruritus is commonly regarded as a hallmark symptom of cholestasis, including in patients with PFICs [3,22,23]. In our study we observed that around a quarter of PFIC1 and PFIC3 patients, and a third of PFIC2 and PFIC4 patients, did not report pruritus. Previous studies indicate that pruritus remains a significant and distressing symptom in PFICs, particularly in PFIC1 and PFIC2, where it is often severe and nearly universal; PFIC3 patients tend to experience milder pruritus, while PFIC4 and PFIC5 patients have less comprehensive data but include pruritus as a symptom for most patients [13,24,25]. However, pruritus may be underreported in preverbal children, leading to potential underestimation of its prevalence [26]. This underscores the need for increased clinician awareness of pruritus to ensure timely and accurate diagnosis and management.

Splenomegaly and portal hypertension were less frequent in PFIC1 patients in our study compared to other PFIC types. This observation aligns with previous findings on the distinct nature of genetic mutations and their impact on liver pathology. PFIC1, caused by mutations in the ATP8B1 gene, leads to a unique progression of liver disease characterized primarily by issues with bile acid transport and liver cell damage, rather than extensive bile duct proliferation and fibrosis. This results in a less frequent progression to cirrhosis and portal hypertension early in the disease course. In PFIC2 and PFIC3 types, mutations in the *ABCB11* and *ABCB4* genes, respectively, often lead to more severe bile secretion defects, increased bile duct proliferation, and quicker progression to liver fibrosis and portal hypertension [22,27].

The higher prevalence of diarrhea in PFIC1 patients compared to other PFIC types can be attributed to the unique pathophysiology associated with mutations in the *ATP8B1* gene. PFIC1 involves defects in the FIC1/Atp8b1 protein, which is crucial for maintaining the balance of bile acids and other lipids in the liver and intestines. This defect leads to steatorrhea and diarrhea due to impaired bile acid reabsorption in the intestines. Additionally, mutations in the *ATP8B1* gene can affect pancreatic function, resulting in exocrine pancreatic insufficiency and subsequent diarrhea. These extrahepatic manifestations are distinct in PFIC1. In contrast, the lower rates of diarrhea observed in PFIC2, PFIC3, and PFIC4 align with their pathophysiological mechanisms, which primarily involve isolated hepatic involvement without significant extrahepatic effects [13,14,16,22].

In our study, 15.8% of PFIC1 and 13.3% of PFIC4 patients developed hearing loss. Previous studies have documented instances of hearing loss in patients with PFIC1 and PFIC4, linking these symptoms to specific genetic mutations. In PFIC1, the *ATP8B1* gene product, the FIC1/Atp8b1 protein, is specifically localized in the stereocilia of cochlear hair cells. The mechanosensory function and integrity of these hair cells critically depend on ATP8B1 activity, which maintains lipid asymmetry in the cellular membranes of stereocilia. Disruptions in this function due to *ATP8B1* mutations can lead to hearing loss [22,28]. Similarly, mutations in the *TJP2* gene, which encodes the tight junction protein 2 (TJP2), can be associated with PFIC4. The TJP2 protein is crucial for maintaining cell-cell adhesion and the integrity of tight junctions in the inner ear, and mutations in *TJP2* can disrupt these structures, leading to hearing loss [29]. Hearing loss in one out of 52 patients with PFIC3 is likely not related to the PFIC disease.

Previous studies have shown that among PFIC types, PFIC3 is characterized by high levels of GGT; in contrast, PFIC1 and PFIC2 typically present with normal or low GGT levels [2,13,14,16,22]. PFIC4 can also exhibit elevated GGT levels, but this is not as consistent as in PFIC3 [17]. However, in our study about half of PFIC1, and PFIC2 patients exhibited elevated GGT levels, though these increases were mild-to-moderate (<2 ULN). The elevations in GGT in PFIC1 and PFIC2 can be attributed to advanced stages of the disease at the time of genetic diagnosis, where prolonged cholestasis and progressive liver damage lead to more significant hepatocytes and biliary injury, resulting in GGT elevation.

Increases in ALT, AST, and ALP, as well as impaired blood coagulation tests, have been well-documented in all types of PFIC. The predominant increase in one or more of these markers among patients with different types of PFIC in our cohort can likely be attributed to the specific set of patients presenting at various stages and severities of the disease at the time of genetic diagnosis in various PFIC groups. This variation reflects the heterogeneous nature of PFIC progression and the liver’s response to ongoing cholestasis and hepatocellular damage.

The literature suggests that PFIC2, PFIC3, and PFIC4 carry an elevated risk for the development of liver tumors [30]. However, we did not observe liver tumors in our study, including among patients who had entered their second decade of life. In our cohort, AFP was tested only in seven patients across different PFIC types during the treatment and follow-up period, with only one patient showing a normal level. The level of AFP that is suspected of hepatocellular carcinoma (HCC) can vary, but generally, an AFP level greater than 400 mcg/L is considered suggestive of HCC in the appropriate clinical context [31]. Moderate elevation of AFP, <400 mcg/L, can be seen in a variety of liver conditions, including chronic hepatitis, cirrhosis, and PFICs [22,23,32]. This level is not specific for HCC but warrants further investigation, as HCC often remains undiagnosed until advanced stages. In many cases, HCC is only identified post-mortem during an autopsy [33]. Therefore, elevated AFP levels, especially when progressively increasing, should prompt additional diagnostic evaluations to identify HCC at an earlier, more treatable stage.

Anemia in PFIC patients can result from various factors related to liver dysfunction and disease impact and usually develops at the advanced stage of the disease. Anemia in PFIC patients arises from several factors: chronic liver disease disrupts iron metabolism and protein production; splenomegaly can lead to hypersplenism, causing excessive red blood cell destruction; nutritional deficiencies due to malabsorption further contribute to anemia; increased bleeding risk through reducing clotting factor synthesis can lead to chronic blood loss; chronic inflammation can suppress bone marrow function, reducing red blood cell production [2,3,14,22,23,34]. Additionally, frequent blood draws during hospital stays exacerbate the condition, particularly in children [35]. A significant majority of PFIC patients in our cohort (84.2%-89.3%) presented with anemia, indicating that most of the patients received their PFIC genetic diagnosis at the advanced stage of the disease.

A common feature among all PFIC types in our cohort was their poor prognosis, with a low cumulative survival rate of 20% (95% CI 5-41%) over 20 years. Consistent with previous reports, untreated PFIC1 and PFIC3 typically progress to end-stage liver disease and death within 10-20 years, with an even worse prognosis in cases of PFIC2 [2,16,36]. A review of 10 studies [37] showed mortality rates ranging up to 87%, with the median age at death being 4 years; survival rates for patients who did not undergo biliary surgery or LT dropped to 50% by age 10, with virtually no survivors by age 20 years. This high mortality rate underscores the frequent need for LT in PFIC patients, with transplantation rates as high as 65% [5].

Literature indicates that PFIC1, PFIC2, PFIC4, and PFIC5 are particularly aggressive in their disease course, often rapidly progressing to end-stage liver disease [23]. This aligns with our observations: PFIC3 exhibited a less aggressive course compared to PFIC1, PFIC2, and PFIC4 in the first decade.

Truncated variants, such as frameshift, nonsense, stop gain, splice, and start loss mutations, lead to the production of dysfunctional or absent proteins, exacerbating the pathophysiological mechanisms of PFIC. Consequently, patients with these variants often present with more severe symptoms, faster disease progression, and poorer outcomes compared to those with non-truncated variants, such as missense and substitution mutations [13,24,38–40]. In our cohort, patients with non-truncated variants showed better, though not statistically significant, cumulative survival (Figure 1D). The increased aggressiveness of the disease in patients with truncated variants is also associated with poorer responses to conventional treatments and a higher necessity for LT. These findings underscore the importance of early genetic diagnosis and personalized treatment approaches to improve outcomes for PFIC patients.

Our study acknowledges certain limitations. There is a possibility that patients with a late onset of symptoms, particularly in their second decade, were not captured in our study. The study did not include patients with cholestasis and negative genetic results, which did not allow analysis and comparison of genetic and idiopathic forms of PFIC.

## 4. Materials and Methods

### Study participants

This observational study analyzed pediatric patients from hospitals in Islamabad, Karachi, Lahore, Multan, and Peshawar, Pakistan. Patients presenting with clinical and laboratory features of cholestatic liver disease from January 2019 to January 2023 underwent WGS. We specifically included individuals possessing homozygous or compound heterozygous variants in a set of genes known to be associated with PFICs: *ATP8B1, ABCB11, ABCB4, TJP2, NR1H4, SLC51A, USP53, KIF12, ZFYVE19, MYO5B, SEMA7A*, and *VPS33B*. Informed consent was obtained from all participants (or their parents / legal guardians) involved in the study. Demographic, clinical, and laboratory features at the time of PFIC genetic diagnosis were recorded and further analyzed. The overall prognosis and outcomes were also collected and patients’ survival was analyzed. Patients who were lost to follow-up, with unknown outcomes were not included in the study. The study was approved by the Ethics Committee of Rostock University (Germany), A2022-0072, 25.04 2022.

### Sequencing and data analysis

Whole Genome Sequencing (WGS). For our study, DNA samples were prepared using the TruSeq DNA Nano Library Prep Kit from Illumina. Sequencing was performed on an Illumina platform utilizing the 150 bp paired-end protocol, achieving an average coverage depth of 30x for the nuclear genome. The alignment of raw reads to the reference genome GRCH38 and the calling of variants, including single nucleotide substitutions (SNVs), small insertions/deletions (Indels), and structural variants (SVs), were conducted using DRAGEN (version 3.10.4, Illumina). Annotation of SNVs and indels was carried out by Varvis (Limbus Medical Technologies GmbH; https://www.limbus-medtec.com/), while structural variants were annotated using ANNOTSV3.1. All genetic variants were described according to the Human Genome Variation Society (HGVS) recommendations (www.hgvs.org).

### Variant evaluation and interpretation

We considered only high-quality variants with a minimum of 9 reads and an alternate allele frequency of at least 0.3%. Candidate variants underwent evaluation for their pathogenicity and causality using a 5-tier classification: pathogenic (P), likely pathogenic (LP), variants of uncertain significance (VUS), likely benign (LB), and benign (B). Our analysis was restricted to genes with clear associations with the participants’ phenotypes, using Human Phenotype Ontology nomenclature (HPO) (https://hpo.jax.org/app/). Factors considered included allele frequency in control databases (gnomAD), in silico pathogenicity predictions, potential protein impacts, variant type-disease mechanisms, familial segregation, and external evidence from OMIM (https://www.omim.org/), ClinVar (https://www.ncbi.nlm.nih.gov/clinvar/), and MasterMind (https://mastermind.genomenon.com/), along with genotype-phenotype correlations.

### Statistics

Statistical analyses were performed using SPSS version 26.0 (IBM Corp., Armonk, NY, USA). Data are presented as median (range) and median (25-75 percentile) for continuous variables, and as frequencies and percentages for categorical variables. Comparisons between groups were conducted using the Chi-square test for categorical variables and one-way analysis of variance (ANOVA) for continuous variables. Post-hoc pairwise comparisons were performed using Dunn’s test with Bonferroni correction for multiple comparisons. Survival rates were calculated using the Kaplan-Meier method, and differences between survival curves were assessed using the log-rank test. A p-value of less than 0.05 was considered statistically significant.

## 5. Conclusions

This study represents the first comprehensive analysis of PFIC in Pakistani children, providing valuable insights into the genetic and phenotypic diversity of this condition. All PFIC types demonstrated significant clinical and laboratory overlap, making differentiation based on these results challenging. WGS proved to be a simple diagnostic tool, crucial for accurate diagnosis.

The clinical presentations, laboratory findings, and outcomes in our study largely align with those reported previously. However, we identified some distinguishing features. In our cohort, PFIC3 emerged as the most prevalent type, followed by PFIC2, PFIC1, PFIC4, and PFIC5. Increased GGT in half of the PFIC1 and PFIC2 patients, as well as anemia in nearly all patients with all PFIC types at the time of genetic diagnosis, can be attributed to delayed PFIC genetic diagnosis in Pakistani children. This highlights the need for the implementation of genetic testing protocols.

The decrease in the proportion of patients with pruritus and the absence of liver tumors in the large cohort may indicate the need for the implementation of additional protocols for pruritus diagnosis in non-verbal children, as well as the implementation of a screening and monitoring system for liver tumors in these patients in Pakistan.

Patients with PFIC1, PFIC2, PFIC4, and/or truncated variants exhibited poorer outcomes, underscoring the importance of early genetic diagnosis and personalized treatment approaches, early consideration for LT in particular, to improve outcomes.

## Data Availability

The variant from this study have been submitted to the NCBI ClinVar database (http://www.clinvar.com/) under accession numbers SCV004232458 - SCV004232527

## Author Contributions

We declare that this work was done by the authors named in this article and all liabilities pertaining to claims relating to the content of this article will be borne by the authors. HAC, AS, ZF, and AR conceived the idea and drafted and revised the manuscript. MAA, MNA, NW, KR, and VS performed participants’ enrolment, clinical evaluation, and sampling. AS, VS, and AM genomic data analysis. AS, AR, VS, and NW manuscript writing.

## Institutional Review Board Statement

The study was approved by the Ethics Committee of Rostock University (Germany), A2022-0072, 25.04 2022.

## Informed Consent Statement

Written informed consent for participation in the study and publication of this paper was obtained from all participants or their parents/legal guardians.

## Data Availability Statement

The genetic variant from this study have been submitted to the NCBI ClinVar database (http://www.clinvar.com/) under accession numbers SCV004232458 - SCV004232527.

## Acknowledgments

We thank the contributions of the administration and staff of the Children Hospital (Lahore), Pakistan Institute of Medical Sciences (Islamabad), Children Hospital and ICH (Multan), Town Women and Children Hospital (Peshawar), and Liaquat National Hospital (Karachi).

---

**Table S1.**
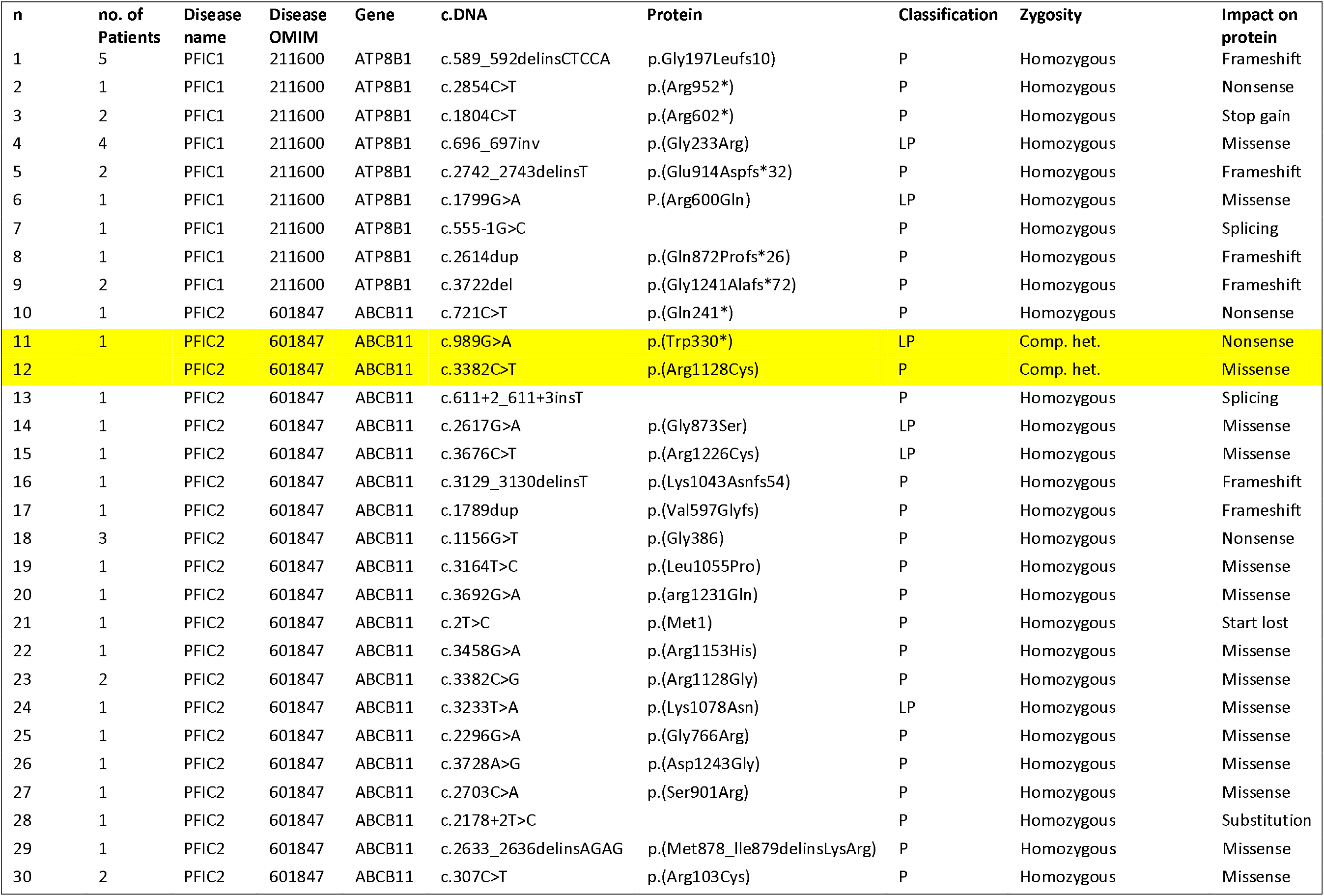
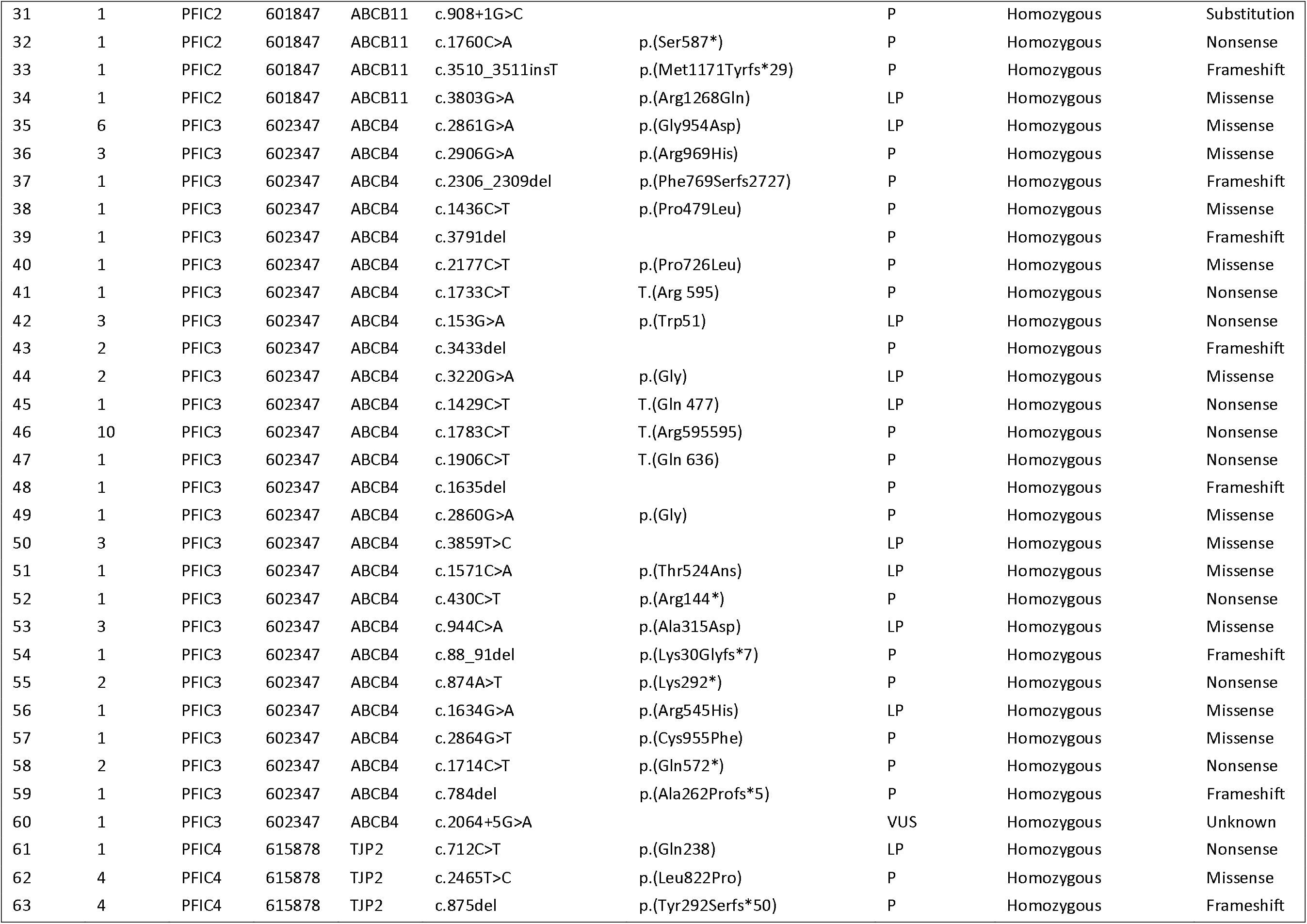
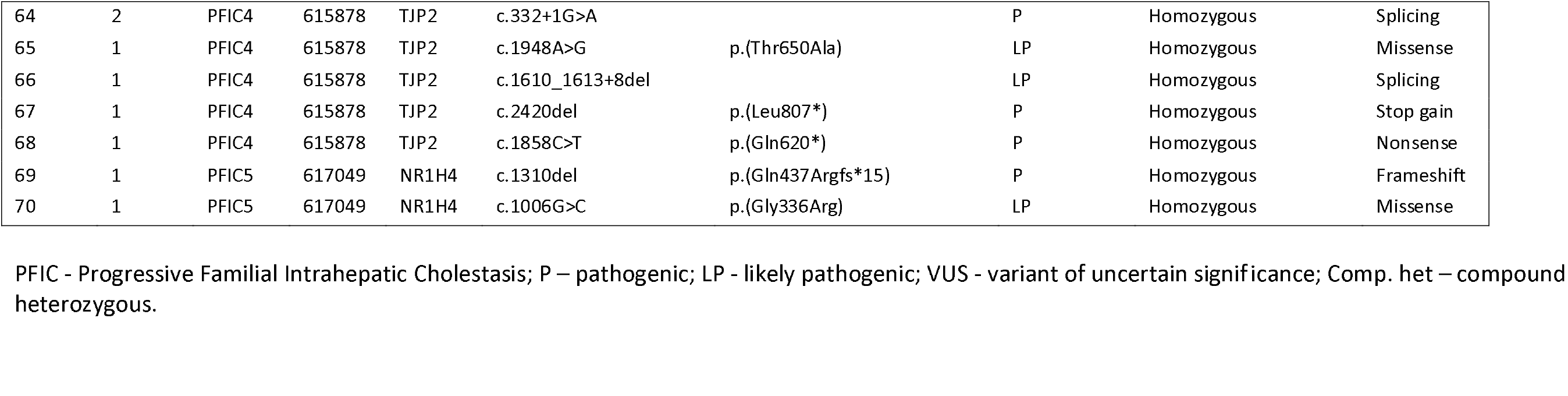
PFIC variants.

## References

1. Gunaydin, M.; Bozkurter Cil, A.T. Progressive Familial Intrahepatic Cholestasis: Diagnosis, Management, and Treatment. Hepatic Med. Evid. Res. 2018, 10, 95–104, doi:10.2147/HMER.S137209.

2. Srivastava, A. Progressive Familial Intrahepatic Cholestasis. J. Clin. Exp. Hepatol. 2014, 4, 25–36, doi:10.1016/j.jceh.2013.10.005.

3. Siddiqi, I.; Tadi, P. Progressive Familial Intrahepatic Cholestasis. In StatPearls; StatPearls Publishing: Treasure Island (FL), 2023.

4. Amberger, J.S.; Hamosh, A. Searching Online Mendelian Inheritance in Man (OMIM): A Knowledgebase of Human Genes and Genetic Phenotypes. Curr. Protoc. Bioinforma. 2017, 58, 1.2.1-1.2.12, doi:10.1002/cpbi.27.

5. Alsohaibani, F.I.; Peedikayil, M.C.; Alfadley, A.F.; Aboueissa, M.K.; Abaalkhail, F.A.; Alqahtani, S.A. Progressive Familial Intrahepatic Cholestasis: A Descriptive Study in a Tertiary Care Center. Int. J. Hepatol. 2023, 2023, 1–8, doi:10.1155/2023/1960152.

6. WHO Multicentre Growth Reference Study Group WHO Child Growth Standards Based on Length/Height, Weight and Age. Acta Paediatr. Oslo Nor. 1992 Suppl. 2006, 450, 76–85, doi:10.1111/j.1651-2227.2006.tb02378.x.

7. Konrad-Martin, D.; Poling, G.L.; Dreisbach, L.E.; Reavis, K.M.; McMillan, G.P.; Lapsley Miller, J.A.; Marshall, L. Serial Monitoring of Otoacoustic Emissions in Clinical Trials. Otol. Neurotol. Off. Publ. Am. Otol. Soc. Am. Neurotol. Soc. Eur. Acad. Otol. Neurotol. 2016, 37, e286–294, doi:10.1097/MAO.0000000000001134.

8. Davit-Spraul, A.; Fabre, M.; Branchereau, S.; Baussan, C.; Gonzales, E.; Stieger, B.; Bernard, O.; Jacquemin, E. ATP8B1 and ABCB11 Analysis in 62 Children with Normal Gamma-Glutamyl Transferase Progressive Familial Intrahepatic Cholestasis (PFIC): Phenotypic Differences between PFIC1 and PFIC2 and Natural History. Hepatol. Baltim. Md 2010, 51, 1645–1655, doi:10.1002/hep.23539.

9. Müllenbach, R.; Lammert, F. An Update on Genetic Analysis of Cholestatic Liver Diseases: Digging Deeper. Dig. Dis. Basel Switz. 2011, 29, 72–77, doi:10.1159/000324137.

10. Sharma, A.; Poddar, U.; Agnihotry, S.; Phadke, S.R.; Yachha, S.K.; Aggarwal, R. Spectrum of Genomic Variations in Indian Patients with Progressive Familial Intrahepatic Cholestasis. BMC Gastroenterol. 2018, 18, 107, doi:10.1186/s12876-018-0835-6.

11. Klomp, L.W.J.; Vargas, J.C.; van Mil, S.W.C.; Pawlikowska, L.; Strautnieks, S.S.; van Eijk, M.J.T.; Juijn, J.A.; Pabón-Peña, C.; Smith, L.B.; DeYoung, J.A.; et al. Characterization of Mutations in ATP8B1 Associated with Hereditary Cholestasis. Hepatol. Baltim. Md 2004, 40, 27–38, doi:10.1002/hep.20285.

12. Lang, T.; Haberl, M.; Jung, D.; Drescher, A.; Schlagenhaufer, R.; Keil, A.; Mornhinweg, E.; Stieger, B.; Kullak-Ublick, G.A.; Kerb, R. Genetic Variability, Haplotype Structures, and Ethnic Diversity of Hepatic Transporters MDR3 (ABCB4) and Bile Salt Export Pump (ABCB11). Drug Metab. Dispos. Biol. Fate Chem. 2006, 34, 1582–1599, doi:10.1124/dmd.105.008854.

13. Alam, S.; Lal, B.B. Recent Updates on Progressive Familial Intrahepatic Cholestasis Types 1, 2 and 3: Outcome and Therapeutic Strategies. World J. Hepatol. 2022, 14, 98–118, doi:10.4254/wjh.v14.i1.98.

14. Baker, A.; Kerkar, N.; Todorova, L.; Kamath, B.M.; Houwen, R.H.J. Systematic Review of Progressive Familial Intrahepatic Cholestasis. Clin. Res. Hepatol. Gastroenterol. 2019, 43, 20–36, doi:10.1016/j.clinre.2018.07.010.

15. Riaz, M.; Tiller, J.; Ajmal, M.; Azam, M.; Qamar, R.; Lacaze, P. Implementation of Public Health Genomics in Pakistan. Eur. J. Hum. Genet. EJHG 2019, 27, 1485–1492, doi:10.1038/s41431-019-0428-z.

16. Vitale, G.; Gitto, S.; Vukotic, R.; Raimondi, F.; Andreone, P. Familial Intrahepatic Cholestasis: New and Wide Perspectives. Dig. Liver Dis. Off. J. Ital. Soc. Gastroenterol. Ital. Assoc. Study Liver 2019, 51, 922–933, doi:10.1016/j.dld.2019.04.013.

17. Sambrotta, M.; Thompson, R.J. Mutations in TJP2, Encoding Zona Occludens 2, and Liver Disease. Tissue Barriers 2015, 3, e1026537, doi:10.1080/21688370.2015.1026537.

18. González-Mariscal, L.; Gallego-Gutiérrez, H.; González-González, L.; Hernández-Guzmán, C. ZO-2 Is a Master Regulator of Gene Expression, Cell Proliferation, Cytoarchitecture, and Cell Size. Int. J. Mol. Sci. 2019, 20, 4128, doi:10.3390/ijms20174128.

19. Smit, J.J.; Schinkel, A.H.; Oude Elferink, R.P.; Groen, A.K.; Wagenaar, E.; van Deemter, L.; Mol, C.A.; Ottenhoff, R.; van der Lugt, N.M.; van Roon, M.A. Homozygous Disruption of the Murine Mdr2 P-Glycoprotein Gene Leads to a Complete Absence of Phospholipid from Bile and to Liver Disease. Cell 1993, 75, 451–462, doi:10.1016/0092-8674(93)90380-9.

20. Popov, Y.; Patsenker, E.; Fickert, P.; Trauner, M.; Schuppan, D. Mdr2 (Abcb4)-/-Mice Spontaneously Develop Severe Biliary Fibrosis via Massive Dysregulation of pro- and Antifibrogenic Genes. J. Hepatol. 2005, 43, 1045–1054, doi:10.1016/j.jhep.2005.06.025.

21. Davit-Spraul, A.; Gonzales, E.; Baussan, C.; Jacquemin, E. Progressive Familial Intrahepatic Cholestasis. Orphanet J. Rare Dis. 2009, 4, 1, doi:10.1186/1750-1172-4-1.

22. Amirneni, S.; Haep, N.; Gad, M.A.; Soto-Gutierrez, A.; Squires, J.E.; Florentino, R.M. Molecular Overview of Progressive Familial Intrahepatic Cholestasis. World J. Gastroenterol. 2020, 26, 7470–7484, doi:10.3748/wjg.v26.i47.7470.

23. Amendola, M.; Squires, J.E. Pediatric Genetic Cholestatic Liver Disease Overview. In GeneReviews®; Adam, M.P., Mirzaa, G.M., Pagon, R.A., Wallace, S.E., Bean, L.J., Gripp, K.W., Amemiya, A., Eds.; University of Washington, Seattle: Seattle (WA), 1993.

24. Jones-Hughes, T.; Campbell, J.; Crathorne, L. Epidemiology and Burden of Progressive Familial Intrahepatic Cholestasis: A Systematic Review. Orphanet J. Rare Dis. 2021, 16, 255, doi:10.1186/s13023-021-01884-4.

25. Henkel, S.A.; Squires, J.H.; Ayers, M.; Ganoza, A.; Mckiernan, P.; Squires, J.E. Expanding Etiology of Progressive Familial Intrahepatic Cholestasis. World J. Hepatol. 2019, 11, 450–463, doi:10.4254/wjh.v11.i5.450.

26. Satoh, T.; Yokozeki, H.; Murota, H.; Tokura, Y.; Kabashima, K.; Takamori, K.; Shiohara, T.; Morita, E.; Aiba, S.; Aoyama, Y.; et al. 2020 Guidelines for the Diagnosis and Treatment of Cutaneous Pruritus. J. Dermatol. 2021, 48, e399–e413, doi:10.1111/1346-8138.16066.

27. Sharjeel, S.; Abdullah, M. Successful Management of Portal Hypertension with Splenomegaly and Cavernous Transformation of the Portal Vein: A Rare Case Report. J. Surg. Case Rep. 2023, 2023, rjad607, doi:10.1093/jscr/rjad607.

28. Stapelbroek, J.M.; Peters, T.A.; van Beurden, D.H.A.; Curfs, J.H.A.J.; Joosten, A.; Beynon, A.J.; van Leeuwen, B.M.; van der Velden, L.M.; Bull, L.; Oude Elferink, R.P.; et al. ATP8B1 Is Essential for Maintaining Normal Hearing. Proc. Natl. Acad. Sci. U. S. A. 2009, 106, 9709–9714, doi:10.1073/pnas.0807919106.

29. Wang, H.-Y.; Zhao, Y.-L.; Liu, Q.; Yuan, H.; Gao, Y.; Lan, L.; Yu, L.; Wang, D.-Y.; Guan, J.; Wang, Q.-J. Identification of Two Disease-Causing Genes TJP2 and GJB2 in a Chinese Family with Unconditional Autosomal Dominant Nonsyndromic Hereditary Hearing Impairment. Chin. Med. J. (Engl.) 2015, 128, 3345–3351, doi:10.4103/0366-6999.171440.

30. Zhou, S.; Hertel, P.M.; Finegold, M.J.; Wang, L.; Kerkar, N.; Wang, J.; Wong, L.-J.C.; Plon, S.E.; Sambrotta, M.; Foskett, P.; et al. Hepatocellular Carcinoma Associated with Tight-Junction Protein 2 Deficiency. Hepatol. Baltim. Md 2015, 62, 1914–1916, doi:10.1002/hep.27872.

31. Bialecki, E.S.; Di Bisceglie, A.M. Diagnosis of Hepatocellular Carcinoma. HPB 2005, 7, 26–34, doi:10.1080/13651820410024049.

32. Gomez-Ospina, N.; Potter, C.J.; Xiao, R.; Manickam, K.; Kim, M.-S.; Kim, K.H.; Shneider, B.L.; Picarsic, J.L.; Jacobson, T.A.; Zhang, J.; et al. Mutations in the Nuclear Bile Acid Receptor FXR Cause Progressive Familial Intrahepatic Cholestasis. Nat. Commun. 2016, 7, 10713, doi:10.1038/ncomms10713.

33. Kaczynski, J.; Hansson, G.; Wallerstedt, S. Clinical Features in Hepatocellular Carcinoma and the Impact of Autopsy on Diagnosis. A Study of 530 Cases from a Low-Endemicity Area. Hepatogastroenterology. 2005, 52, 1798–1802.

34. Agarwal, S.; Lal, B.B.; Rawat, D.; Rastogi, A.; Bharathy, K.G.S.; Alam, S. Progressive Familial Intrahepatic Cholestasis (PFIC) in Indian Children: Clinical Spectrum and Outcome. J. Clin. Exp. Hepatol. 2016, 6, 203–208, doi:10.1016/j.jceh.2016.05.003.

35. Eaton, K.P.; Levy, K.; Soong, C.; Pahwa, A.K.; Petrilli, C.; Ziemba, J.B.; Cho, H.J.; Alban, R.; Blanck, J.F.; Parsons, A.S. Evidence-Based Guidelines to Eliminate Repetitive Laboratory Testing. JAMA Intern. Med. 2017, 177, 1833–1839, doi:10.1001/jamainternmed.2017.5152.

36. Hori, T.; Nguyen, J.H.; Uemoto, S. Progressive Familial Intrahepatic Cholestasis. Hepatobiliary Pancreat. Dis. Int. HBPD INT 2010, 9, 570–578.

37. Baker, A.; Kerkar, N.; Todorova, L.; Kamath, B.M.; Houwen, R.H.J. Erratum to “Systematic Review of Progressive Familial Intrahepatic Cholestasis” [Clin. Res. Hepatol. Gastroenterol. 43 (2019) 20-36]. Clin. Res. Hepatol. Gastroenterol. 2020, 44, 115, doi:10.1016/j.clinre.2019.12.005.

38. Chen, R.; Yang, F.-X.; Tan, Y.-F.; Deng, M.; Li, H.; Xu, Y.; Ouyang, W.-X.; Song, Y.-Z. Clinical and Genetic Characterization of Pediatric Patients with Progressive Familial Intrahepatic Cholestasis Type 3 (PFIC3): Identification of 14 Novel ABCB4 Variants and Review of the Literatures. Orphanet J. Rare Dis. 2022, 17, 445, doi:10.1186/s13023-022-02597-y.

39. van Wessel, D.B.E.; Thompson, R.J.; Gonzales, E.; Jankowska, I.; Shneider, B.L.; Sokal, E.; Grammatikopoulos, T.; Kadaristiana, A.; Jacquemin, E.; Spraul, A.; et al. Impact of Genotype, Serum Bile Acids, and Surgical Biliary Diversion on Native Liver Survival in FIC1 Deficiency. Hepatol. Baltim. Md 2021, 74, 892–906, doi:10.1002/hep.31787.

40. Lal, B.B.; Alam, S.; Sibal, A.; Kumar, K.; Hosaagrahara Ramakrishna, S.; Shah, V.; Dheivamani, N.; Bavdekar, A.; Nagral, A.; Wadhwa, N.; et al. Genotype Correlates with Clinical Course and Outcome of Children with Tight Junction Protein 2 (TJP2) Deficiency-Related Cholestasis. Hepatol. Baltim. Md 2024, doi:10.1097/HEP.0000000000000828.

