## Supplemental Table 1 (Table S1) for "Clinical Diversity and Outcomes of Progressive Familial Intrahepatic Cholestasis Diagnosed by Whole Genome Sequencing in Pakistani Children"

Table S1. PFIC variants.

| **n** | **no. of Patients** | **Disease name** | **Disease OMIM** | **Gene** | **c.DNA** | **Protein** | **Classification** | **Zygosity** | **Impact on protein** |
| --- | --- | --- | --- | --- | --- | --- | --- | --- | --- |
| 1 | 5 | PFIC1 | 211600 | ATP8B1 | c.589_592delinsCTCCA | p.Gly197Leufs10) | P | Homozygous | Frameshift |
| 2 | 1 | PFIC1 | 211600 | ATP8B1 | c.2854C>T | p.(Arg952*) | P | Homozygous | Nonsense |
| 3 | 2 | PFIC1 | 211600 | ATP8B1 | c.1804C>T | p.(Arg602*) | P | Homozygous | Stop gain |
| 4 | 4 | PFIC1 | 211600 | ATP8B1 | c.696_697inv | p.(Gly233Arg) | LP | Homozygous | Missense |
| 5 | 2 | PFIC1 | 211600 | ATP8B1 | c.2742_2743delinsT | p.(Glu914Aspfs*32) | P | Homozygous | Frameshift |
| 6 | 1 | PFIC1 | 211600 | ATP8B1 | c.1799G>A | P.(Arg600Gln) | LP | Homozygous | Missense |
| 7 | 1 | PFIC1 | 211600 | ATP8B1 | c.555-1G>C |  | P | Homozygous | Splicing |
| 8 | 1 | PFIC1 | 211600 | ATP8B1 | c.2614dup | p.(Gln872Profs*26) | P | Homozygous | Frameshift |
| 9 | 2 | PFIC1 | 211600 | ATP8B1 | c.3722del | p.(Gly1241Alafs*72) | P | Homozygous | Frameshift |
| 10 | 1 | PFIC2 | 601847 | ABCB11 | c.721C>T | p.(Gln241*) | P | Homozygous | Nonsense |
| 11 | 1 | PFIC2 | 601847 | ABCB11 | c.989G>A | p.(Trp330*) | LP | Comp. het. | Nonsense |
| 12 |  | PFIC2 | 601847 | ABCB11 | c.3382C>T | p.(Arg1128Cys) | P | Comp. het. | Missense |
| 13 | 1 | PFIC2 | 601847 | ABCB11 | c.611+2_611+3insT |  | P | Homozygous | Splicing |
| 14 | 1 | PFIC2 | 601847 | ABCB11 | c.2617G>A | p.(Gly873Ser) | LP | Homozygous | Missense |
| 15 | 1 | PFIC2 | 601847 | ABCB11 | c.3676C>T | p.(Arg1226Cys) | LP | Homozygous | Missense |
| 16 | 1 | PFIC2 | 601847 | ABCB11 | c.3129_3130delinsT | p.(Lys1043Asnfs54) | P | Homozygous | Frameshift |
| 17 | 1 | PFIC2 | 601847 | ABCB11 | c.1789dup | p.(Val597Glyfs) | P | Homozygous | Frameshift |
| 18 | 3 | PFIC2 | 601847 | ABCB11 | c.1156G>T | p.(Gly386) | P | Homozygous | Nonsense |
| 19 | 1 | PFIC2 | 601847 | ABCB11 | c.3164T>C | p.(Leu1055Pro) | P | Homozygous | Missense |
| 20 | 1 | PFIC2 | 601847 | ABCB11 | c.3692G>A | p.(arg1231Gln) | P | Homozygous | Missense |
| 21 | 1 | PFIC2 | 601847 | ABCB11 | c.2T>C | p.(Met1) | P | Homozygous | Start lost |
| 22 | 1 | PFIC2 | 601847 | ABCB11 | c.3458G>A | p.(Arg1153His) | P | Homozygous | Missense |
| 23 | 2 | PFIC2 | 601847 | ABCB11 | c.3382C>G | p.(Arg1128Gly) | P | Homozygous | Missense |
| 24 | 1 | PFIC2 | 601847 | ABCB11 | c.3233T>A | p.(Lys1078Asn) | LP | Homozygous | Missense |
| 25 | 1 | PFIC2 | 601847 | ABCB11 | c.2296G>A | p.(Gly766Arg) | P | Homozygous | Missense |
| 26 | 1 | PFIC2 | 601847 | ABCB11 | c.3728A>G | p.(Asp1243Gly) | P | Homozygous | Missense |
| 27 | 1 | PFIC2 | 601847 | ABCB11 | c.2703C>A | p.(Ser901Arg) | P | Homozygous | Missense |
| 28 | 1 | PFIC2 | 601847 | ABCB11 | c.2178+2T>C |  | P | Homozygous | Substitution |
| 29 | 1 | PFIC2 | 601847 | ABCB11 | c.2633_2636delinsAGAG | p.(Met878_lle879delinsLysArg) | P | Homozygous | Missense |
| 30 | 2 | PFIC2 | 601847 | ABCB11 | c.307C>T | p.(Arg103Cys) | P | Homozygous | Missense |
| 31 | 1 | PFIC2 | 601847 | ABCB11 | c.908+1G>C |  | P | Homozygous | Substitution |
| 32 | 1 | PFIC2 | 601847 | ABCB11 | c.1760C>A | p.(Ser587*) | P | Homozygous | Nonsense |
| 33 | 1 | PFIC2 | 601847 | ABCB11 | c.3510_3511insT | p.(Met1171Tyrfs*29) | P | Homozygous | Frameshift |
| 34 | 1 | PFIC2 | 601847 | ABCB11 | c.3803G>A | p.(Arg1268Gln) | LP | Homozygous | Missense |
| 35 | 6 | PFIC3 | 602347 | ABCB4 | c.2861G>A | p.(Gly954Asp) | LP | Homozygous | Missense |
| 36 | 3 | PFIC3 | 602347 | ABCB4 | c.2906G>A | p.(Arg969His) | P | Homozygous | Missense |
| 37 | 1 | PFIC3 | 602347 | ABCB4 | c.2306_2309del | p.(Phe769Serfs2727) | P | Homozygous | Frameshift |
| 38 | 1 | PFIC3 | 602347 | ABCB4 | c.1436C>T | p.(Pro479Leu) | P | Homozygous | Missense |
| 39 | 1 | PFIC3 | 602347 | ABCB4 | c.3791del |  | P | Homozygous | Frameshift |
| 40 | 1 | PFIC3 | 602347 | ABCB4 | c.2177C>T | p.(Pro726Leu) | P | Homozygous | Missense |
| 41 | 1 | PFIC3 | 602347 | ABCB4 | c.1733C>T | T.(Arg 595) | P | Homozygous | Nonsense |
| 42 | 3 | PFIC3 | 602347 | ABCB4 | c.153G>A | p.(Trp51) | LP | Homozygous | Nonsense |
| 43 | 2 | PFIC3 | 602347 | ABCB4 | c.3433del |  | P | Homozygous | Frameshift |
| 44 | 2 | PFIC3 | 602347 | ABCB4 | c.3220G>A | p.(Gly) | LP | Homozygous | Missense |
| 45 | 1 | PFIC3 | 602347 | ABCB4 | c.1429C>T | T.(Gln 477) | LP | Homozygous | Nonsense |
| 46 | 10 | PFIC3 | 602347 | ABCB4 | c.1783C>T | T.(Arg595595) | P | Homozygous | Nonsense |
| 47 | 1 | PFIC3 | 602347 | ABCB4 | c.1906C>T | T.(Gln 636) | P | Homozygous | Nonsense |
| 48 | 1 | PFIC3 | 602347 | ABCB4 | c.1635del |  | P | Homozygous | Frameshift |
| 49 | 1 | PFIC3 | 602347 | ABCB4 | c.2860G>A | p.(Gly) | P | Homozygous | Missense |
| 50 | 3 | PFIC3 | 602347 | ABCB4 | c.3859T>C |  | LP | Homozygous | Missense |
| 51 | 1 | PFIC3 | 602347 | ABCB4 | c.1571C>A | p.(Thr524Ans) | LP | Homozygous | Missense |
| 52 | 1 | PFIC3 | 602347 | ABCB4 | c.430C>T | p.(Arg144*) | P | Homozygous | Nonsense |
| 53 | 3 | PFIC3 | 602347 | ABCB4 | c.944C>A | p.(Ala315Asp) | LP | Homozygous | Missense |
| 54 | 1 | PFIC3 | 602347 | ABCB4 | c.88_91del | p.(Lys30Glyfs*7) | P | Homozygous | Frameshift |
| 55 | 2 | PFIC3 | 602347 | ABCB4 | c.874A>T | p.(Lys292*) | P | Homozygous | Nonsense |
| 56 | 1 | PFIC3 | 602347 | ABCB4 | c.1634G>A | p.(Arg545His) | LP | Homozygous | Missense |
| 57 | 1 | PFIC3 | 602347 | ABCB4 | c.2864G>T | p.(Cys955Phe) | P | Homozygous | Missense |
| 58 | 2 | PFIC3 | 602347 | ABCB4 | c.1714C>T | p.(Gln572*) | P | Homozygous | Nonsense |
| 59 | 1 | PFIC3 | 602347 | ABCB4 | c.784del | p.(Ala262Profs*5) | P | Homozygous | Frameshift |
| 60 | 1 | PFIC3 | 602347 | ABCB4 | c.2064+5G>A |  | VUS | Homozygous | Unknown |
| 61 | 1 | PFIC4 | 615878 | TJP2 | c.712C>T | p.(Gln238) | LP | Homozygous | Nonsense |
| 62 | 4 | PFIC4 | 615878 | TJP2 | c.2465T>C | p.(Leu822Pro) | P | Homozygous | Missense |
| 63 | 4 | PFIC4 | 615878 | TJP2 | c.875del | p.(Tyr292Serfs*50) | P | Homozygous | Frameshift |
| 64 | 2 | PFIC4 | 615878 | TJP2 | c.332+1G>A |  | P | Homozygous | Splicing |
| 65 | 1 | PFIC4 | 615878 | TJP2 | c.1948A>G | p.(Thr650Ala) | LP | Homozygous | Missense |
| 66 | 1 | PFIC4 | 615878 | TJP2 | c.1610_1613+8del |  | LP | Homozygous | Splicing |
| 67 | 1 | PFIC4 | 615878 | TJP2 | c.2420del | p.(Leu807*) | P | Homozygous | Stop gain |
| 68 | 1 | PFIC4 | 615878 | TJP2 | c.1858C>T | p.(Gln620*) | P | Homozygous | Nonsense |
| 69 | 1 | PFIC5 | 617049 | NR1H4 | c.1310del | p.(Gln437Argfs*15) | P | Homozygous | Frameshift |
| 70 | 1 | PFIC5 | 617049 | NR1H4 | c.1006G>C | p.(Gly336Arg) | LP | Homozygous | Missense |

PFIC - Progressive Familial Intrahepatic Cholestasis; P – pathogenic; LP - likely pathogenic; VUS - variant of uncertain significance; Comp. het – compound heterozygous.
